# Orbital Incidentaloma: Clinicoradiologic Characteristics and Management Considerations

**DOI:** 10.1101/2025.03.09.25323627

**Authors:** Yeong A Choi, Min Kyu Yang, Ho-Seok Sa

## Abstract

**Purpose:** To investigate the clinical and radiologic characteristics, treatment outcomes, and management strategies for orbital incidentalomas.

**Materials and methods:** We retrospectively reviewed 43 patients with orbital tumors incidentally identified through imaging conducted for unrelated reasons between March 2015 and July 2023. Data on imaging indications, clinicoradiologic features, diagnoses, treatments, and outcomes were analyzed. Patients were categorized into the surgery and observation groups, and their clinical and radiological characteristics were compared.

**Results:** Among the 43 cases, 20 patients (46.5%) were male, with a mean age of 57.1 years and a mean follow-up of 2.77 years. Initial imaging was most commonly conducted for health check-ups (48.8%), headaches (27.9%), or dizziness (14.0%). Common clinical signs included proptosis (41.9%), peripheral diplopia (21.4%), and hypoglobus (9.3%). Benign lesions, such as cavernous venous malformations (55.8%) and schwannomas (27.9%), predominated, with one case of lymphoma. Patients in the surgery group (n=14, 32.6%) were significantly more likely to present with clinical signs, including proptosis, diplopia, and hypoglobus (all p<0.05), and have anterior tumor locations (71.4% vs. 13.8%, p=0.001) compared to the observation group (n=29, 67.4%). Surgical removal was performed without complications in all cases. In the observation group, tumor size remained stable in 96.6% of cases, with no functional deficits identified throughout follow-up.

**Conclusion:** Orbital incidentalomas are often detected during health check-ups or neuroimaging and may exhibit mild proptosis or diplopia. Observation is recommended for asymptomatic posterior orbital lesions, while surgical removal is indicated for anterior lesions with significant clinical signs.

## INTRODUCTION

The increasing utilization of imaging modalities for the central nervous system and orbit has led to a rise in the incidental detection of lesions (1). The term “incidentaloma” was originally coined to describe an adrenal mass discovered incidentally during imaging for reasons unrelated to suspected adrenal disease (2). These incidental lesions can be found in various organs, including the pituitary, thyroid, and salivary glands, paranasal sinuses, pancreas, kidneys, and lungs (3, 4). However, research on orbital incidentalomas remains relatively limited.

An orbital incidentaloma is defined as a lesion within the orbit detected incidentally through imaging performed for reasons unrelated to a suspected orbital mass. While primary orbital mass lesions are relatively rare, with an incidence of 2.02 per million person-years, incidentalomas in the orbit have been increasingly identified due to the widespread use of various imaging modalities (5). Despite their growing recognition, the features and diagnoses of orbital incidentalomas based on radiologic or pathologic findings remain unclear.

Orbital incidentalomas can cause unnecessary anxiety and may lead to unnecessary diagnostic procedures. Some lesions, however, may represent serious conditions, such as primary malignancies, metastatic tumors, or vascular lesions with haemorrhage (6). While Melmed et al. (7) demonstrated that some pituitary incidentalomas remain stable without significant growth over years or even decades, necessitating regular magnetic resonance imaging (MRI) follow-up only in cases of functioning adenomas or specific clinical circumstances, the literature on the long-term follow-up and prognosis of orbital incidentalomas is limited. Therefore, appropriate practice guidelines are warranted.

This study aims to assess the reasons for initial imaging, clinical characteristics, radiological features, treatments, and outcomes of orbital incidentalomas. Furthermore, it seeks to investigate treatment strategies and long-term prognosis for these lesions through extended follow-up.

## MATERIALS AND METHODS

We conducted a retrospective analysis of orbital incidentalomas referred to the Department of Ophthalmology at Asan Medical Center, Seoul, Korea, between March 2015 and July 2023. A single senior author (H-SS) evaluated and treated all the patients. This research was approved by the Institutional Review Board (IRB) of Asan Medical Center (Seoul, Korea; IRB number: 2024-1462) and complied with the Health Insurance Portability and Accountability Act. The research adhered to the principles of the Declaration of Helsinki.

The inclusion criteria for this study comprised patients with orbital tumors incidentally detected during imaging studies conducted for reasons unrelated to suspected orbital disease. Exclusion criteria included patients presenting with ophthalmic symptoms at initial presentation, those without follow-up imaging, and those with a follow-up period below 6 months. Clinical data collected included demographics, reasons for undergoing imaging, symptoms and signs, ophthalmological findings, radiological characteristics, diagnoses (radiologic or pathological), treatment modalities, and outcomes.

We analyzed the initial imaging findings of all patients and performed additional orbital MRI when the initial imaging lacked sufficient detail for a comprehensive assessment. Tumor characteristics assessed included size, location, shape (categorized as oval, lobulated, multi-lobulated, round, elongated, or irregular), and T2 signal intensities. Tumor size was determined by measuring the maximum diameter of the largest mass. Management plans were established based on clinical assessments by surgeons and patients’ individual needs. Surgical intervention was indicated for cases presenting with eyeball displacement (e.g., proptosis or hypoglobus), suspected malignancy requiring biopsy, discomfort due to peripheral diplopia or visual disturbance, or when the patient’s preference for removal and the surgeon’s assessment of surgical feasibility were satisfied. We classified patients into two groups: the surgery group, comprising those who underwent surgical excision of the tumors, and the observation group, comprising those monitored with regular imaging without surgery. Clinical and radiological characteristics were compared between these two groups.

Statistical analysis was performed using IBM SPSS Statistics (Version 26.0; IBM Corp., Armonk, NY, USA). Ordinal data and continuous variables are presented as medians with their respective minimum-maximum ranges. Logistic regression models were employed to assess the associations between tumor size, location, and the necessity for surgical resection. The Chi-square (χ^2^) test and analysis of variance (ANOVA) (for normally distributed data) were used to compare categorical data. A p-value < 0.05 was considered statistically significant.

## RESULTS

Forty-three patients were retrospectively assessed. Table 1 summarizes the clinical data of orbital incidentalomas. The median age was 57.1 years (3–75 years), and 20 patients (46.5%) were male. The median follow-up time was 2.77 years (0.8–10.6 years). The primary reasons for performing imaging tests included health check-ups (21/43, 48.8%), headaches (12/43, 27.9%), dizziness (6/43, 14.0%), blunt head trauma (3/43, 7.0%), and general weakness (1/43, 2.3%). At presentation, patients with orbital incidentalomas did not report any ocular symptoms; however, ophthalmological examination revealed various signs, including proptosis (≥ 2 mm) (18/43, 41.9%), peripheral diplopia (beyond 30°) (9/43, 21.4%), hypoglobus (≥ 2 mm) (4/43, 9.3%), palpable mass (2/43, 4.7%), visual field defects (2/43, 4.7%), upper eyelid oedema (1/43, 2.3%), optic disc oedema (1/43, 2.3%), and pupil defects (1/43, 2.3%).

**Table 1.**
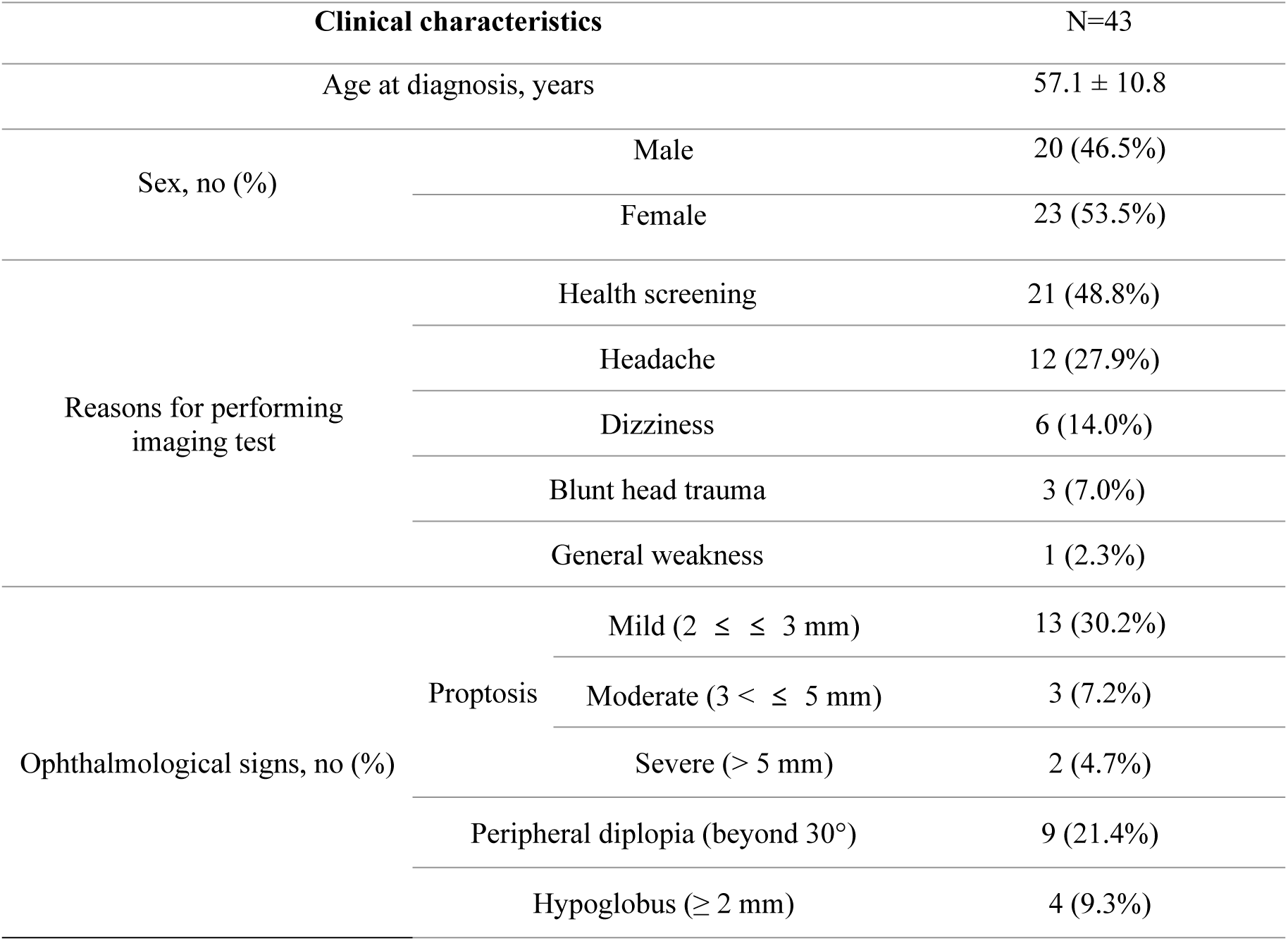
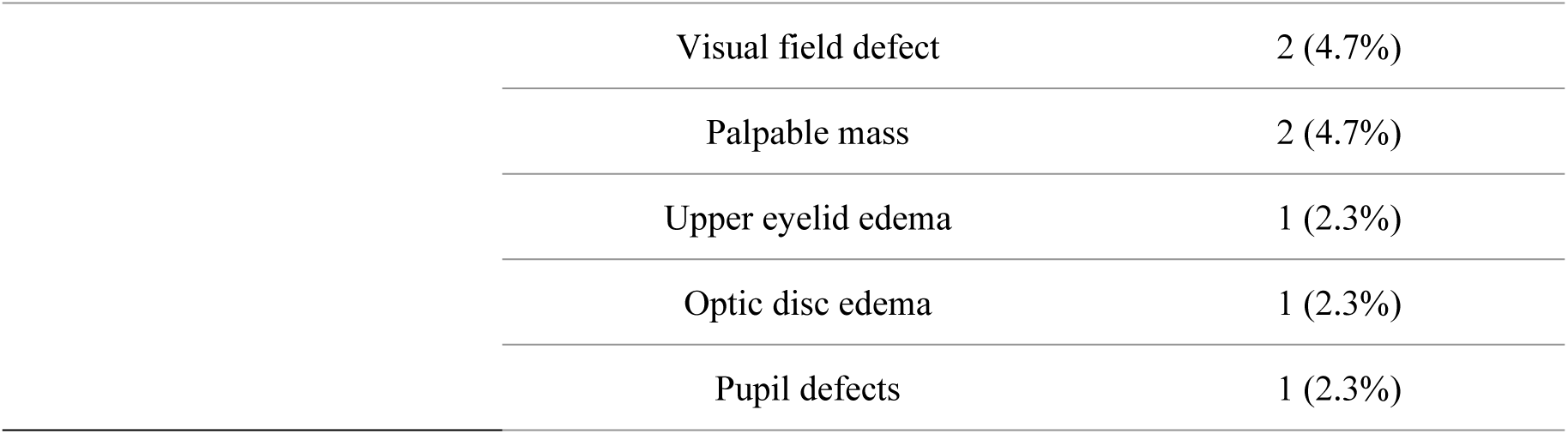
Clinical characteristics of orbital incidentalomas.

Table 2 summarizes the radiologic characteristics of orbital incidentalomas. Among the 43 cases, the radiologic or pathological diagnoses included cavernous venous malformation (CVM) (24/43, 55.8%), schwannoma (12/43, 27.9%), dermoid cyst (2/43, 4.5%), and lymphoma, lymphangioma, fibrous dysplasia, optic glioma, and meningioma (1/43, 2.3% each). The mean tumor size was 16.3 mm (5–40 mm). Tumor locations were more commonly posterior (30/43, 69.8%) than anterior (13/43, 30.2%), and intraconal tumors (31/43, 72.1%) were more frequent than extraconal ones (12/43, 27.9%). By orbital quadrants, superolateral lesions (13/43, 30.2%) were the most common, followed by superomedial lesions (12/43, 27.9%). The most common shape of orbital incidentalomas was oval (26/43, 60.5%), followed by lobulated (6/43, 14.0%). Most tumors were well-circumscribed, with three exceptions: lymphoma, fibrous dysplasia, and optic glioma, which exhibited irregular shapes.

**Table 2.**
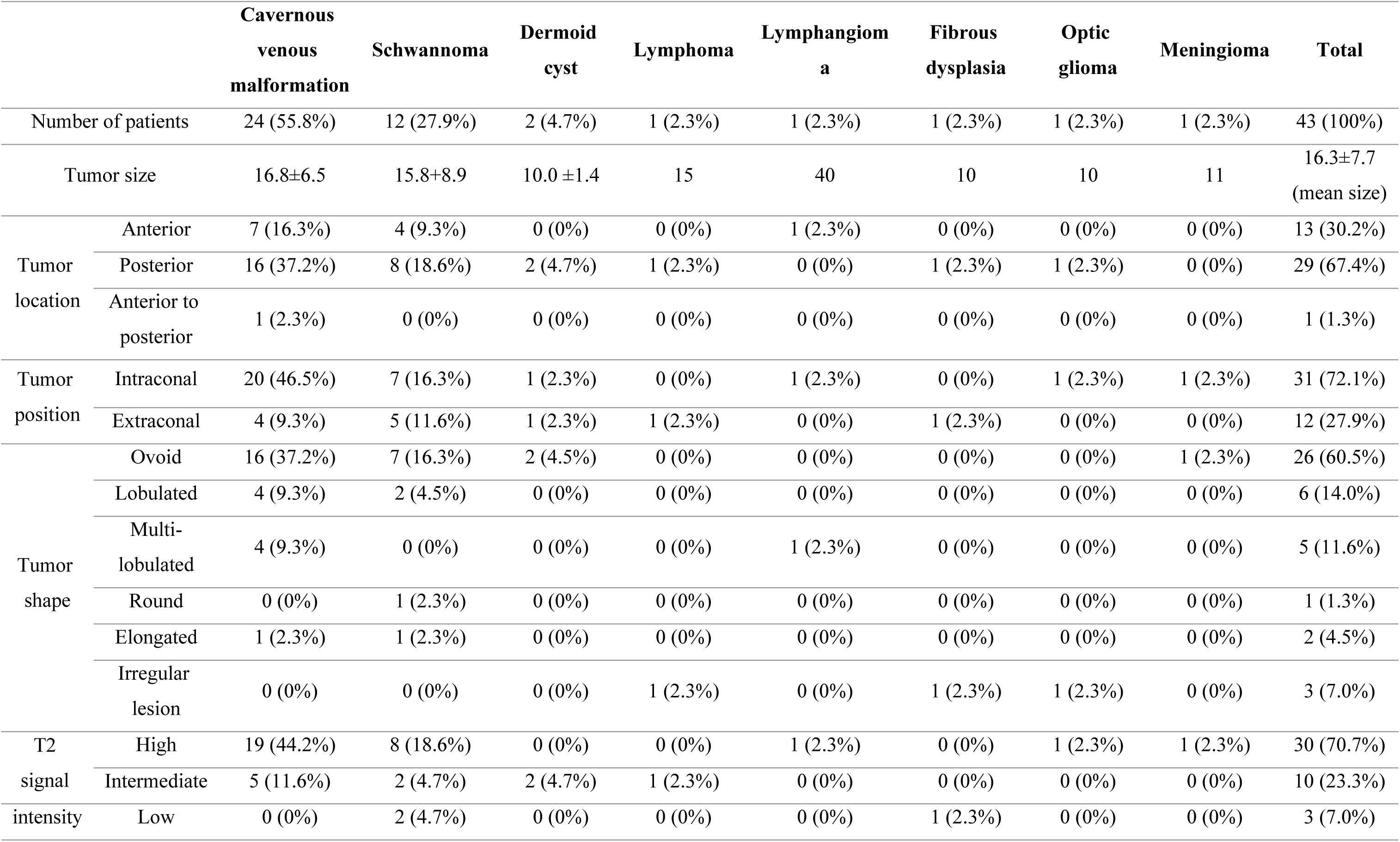
Radiologic characteristics by diagnosis of orbital incidentalomas.

In the surgery group (14/43, 32.6%), surgery included tumor excision (13/43, 30.2%) or incisional biopsy for suspected malignancy (1/43, 2.3%). Indications for surgery included proptosis (≥ 2 mm) (8/14, 57.1%), hypoglobus (2/14, 14.3%), peripheral diplopia (2/14, 14.3%), biopsy for suspected malignancy (1/14, 7.1%), and a palpable mass (1/14, 7.1%). No surgery-related complications were observed. Pathological diagnoses included CVM (n = 7), schwannoma (n = 5), lymphoma (n = 1), and fibrous dysplasia (n = 1). One case initially diagnosed radiologically as CVM was later confirmed pathologically as schwannoma. The patient with biopsy-confirmed lymphoma showed no systemic involvement, underwent radiation therapy, and achieved complete remission without relapse. Among the 12 cases of surgical excision for CVM or schwannoma, no recurrences were observed during a median follow-up of 2.53 years (0.8–3.2 years).

For the observation group (29/43, 67.4%), all patients were monitored during a median follow-up of 2.77 years (1.4–6.2 years) following the initial diagnosis. Ophthalmological examinations and orbital imaging were performed at each visit to evaluate functional status and tumor size changes. Reasons for observation without surgery included concern for complications due to tumor proximity to the optic nerve (13 cases, 44.9%), no symptoms or signs (10 cases, 34.5%), posterior tumor location with challenging surgical access (8 cases, 27.6%), and patient refusal of feasible surgery (2 cases, 6.9%). During follow-up, most patients (26/29, 89.7%) exhibited no changes in tumor size or development of new functional deficits (Fig 1). Tumor size changes were observed in three patients (10.3%): one CVM case exhibited 12% growth (from 17 to 19 mm) over 48 months, while two cases showed tumor reduction— a 25% decrease (from 40 to 30 mm) in lymphangioma at 38.3 months, and a 20% decrease (from 20 to 16 mm) in schwannoma at 22.4 months.

**Fig 1.**
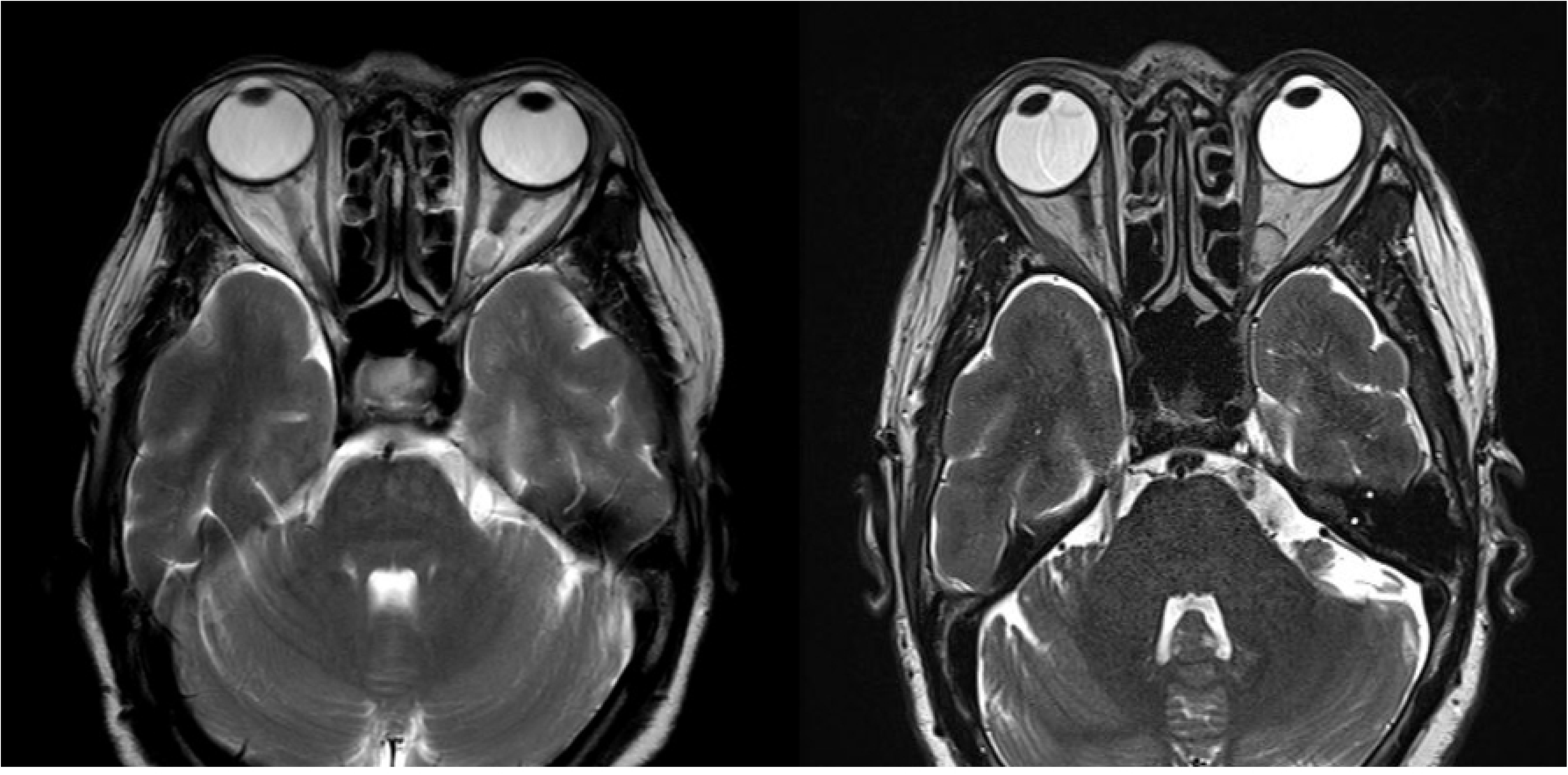
A) MRI at initial presentation supporting the diagnosis of orbital cavernous venous malformation. B) MRI after 2 years showing no mass growth during the observation period.

Table 3 presents and compares the clinical and radiological characteristics of the observation and surgery groups. For ophthalmological signs, proptosis, peripheral diplopia, hypoglobus, and palpable mass were significantly more common in the surgery group than in the observation group (p < 0.001, p = 0.014, p = 0.003, and p = 0.037, respectively). The mean tumor size was larger in the surgery group (19.36 mm) compared to the observation group (14.76 mm), with no significant difference (p = 0.072). Additionally, tumor location differed significantly between the two groups: 71.4% of tumors in the surgery group were located in the anterior orbit, while 86.2% of tumors in the observation group were located in the posterior orbit (p = 0.001). No significant differences were observed in tumor position (intraconal vs extraconal) or shape between the two groups.

**Table 3.**
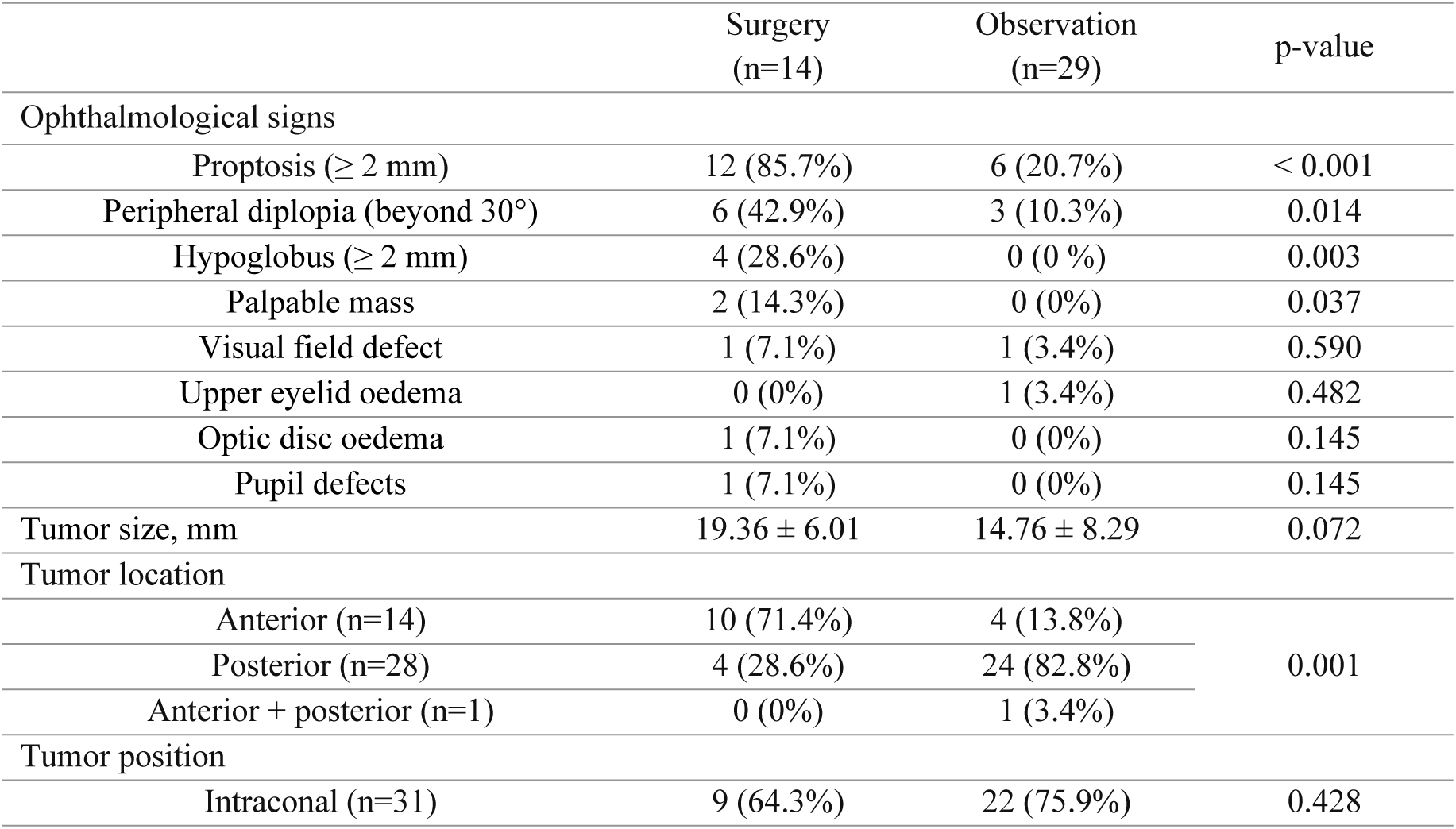
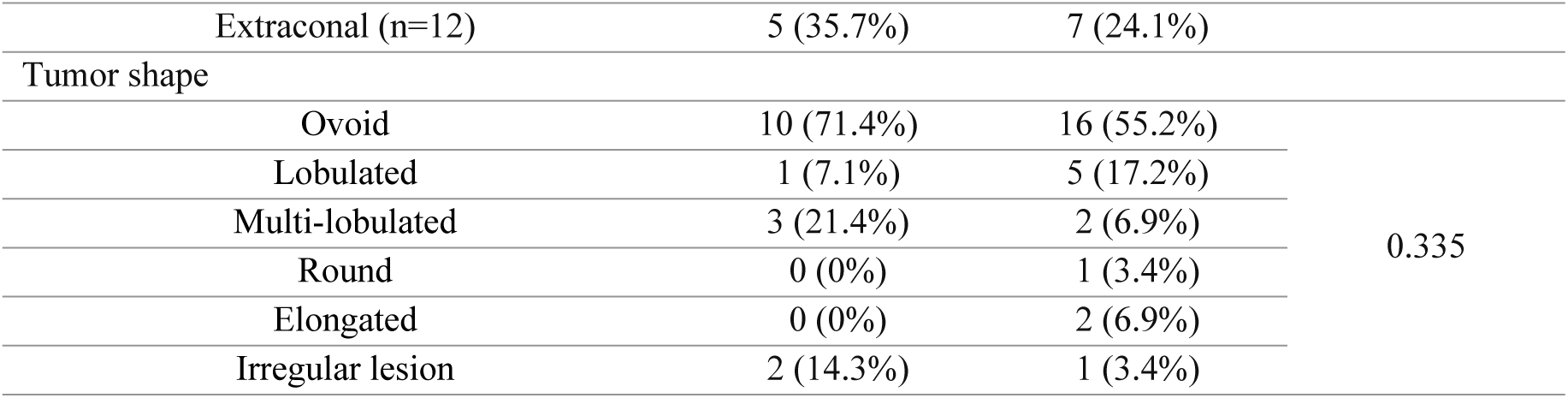
Comparison of clinical and radiological characteristics between observation and surgery groups of orbital incidentalomas.

## DISCUSSION

This study investigated the clinical and radiologic characteristics and management outcomes of orbital incidentalomas in 43 patients, with a median follow-up of 2.8 years. Our findings revealed that orbital incidentalomas were typically identified during imaging performed for health check-ups (49%) or neurological evaluations (42%). The most common diagnoses were CVM (56%) and schwannoma (28%), although a malignant tumor, such as lymphoma, was also detected. None of the patients reported subjective symptoms before diagnosis; however, ophthalmological examinations frequently revealed various signs, such as proptosis (42%), peripheral diplopia (21%), and hypoglobus (10%). We found that most orbital incidentalomas remained stable in size, with only one case (3%) showing minimal growth on follow-up imaging. For incidentalomas in the posterior orbit without significant ophthalmological signs or functional deficits, conservative observation appears to be a reasonable and safe management strategy. In contrast, surgical removal may be considered for anterior orbital lesions due to the typically low risk of complications.

In our study, orbital incidentalomas were predominantly detected during routine health check-ups or neuroimaging studies conducted for unrelated conditions. With improved access to healthcare and increasing use of such tests, the detection of orbital incidentalomas is expected to rise, highlighting the need for a better understanding of their characteristics, clinical courses, and management strategies. Notably, CVMs and schwannomas were our study’s most commonly identified incidentalomas, consistent with previous reports (8). CVMs are recognised as the most common benign orbital tumors in adults, while schwannomas, arising from peripheral nerves, may also occur without obvious clinical symptoms (8, 9). The predominance of these lesions reflects not only their high prevalence but also their characteristic growth patterns and benign nature, often allowing them to remain asymptomatic until discovered incidentally. Radiologically, these lesions typically appear as benign, ovoid or elongated masses, predominantly in the posterior intraconal space, with high T2-weighted signal intensity (10, 11). Previous studies have emphasized the accuracy of radiological diagnosis for CVMs and schwannomas, often eliminating the need for diagnostic surgery (10, 12, 13). Advanced imaging techniques, such as diffusion-weighted MRI, provide valuable information, particularly in distinguishing benign from potentially aggressive pathologies. These findings underscore the critical role of imaging in guiding clinical decisions and supporting conservative management, especially for asymptomatic cases with benign characteristics.

The anatomical location of orbital lesions significantly influences the clinical symptoms and signs (14). In our study, the most common clinical sign was proptosis (42%), followed by peripheral diplopia (21%) and hypoglobus (9%). Notably, approximately 70% of orbital incidentalomas were in the posterior orbit and intraconal space. Small masses in the posterior orbit are often asymptomatic unless apical lesions cause compressive optic neuropathy. However, as these masses increase in size, posterior and intraconal masses can exert pressure along the visual axis, leading to forward displacement of the globe and manifesting as proptosis. This explains why orbital incidentalomas are more commonly found in the posterior orbit, often presenting with gradual and mild proptosis that patients may not initially notice. In contrast, anteriorly located lesions, particularly those near the globe or extraocular muscle, tend to displace ocular structures to the opposite side of the lesion, resulting in hypoglobus, peripheral diplopia, or the presence of a palpable mass. Although patients did not initially recognise these clinical signs before incidental detection, many became more aware of and concerned about these symptoms after diagnosis compared to mild proptosis. This difference appeared to influence patients’ decisions regarding surgical management.

Additionally, although observed in a small number of cases, some patients exhibited visual field defects (5%), optic disc oedema (2%), or pupil defects (2%). These findings highlight the potential for orbital incidentalomas to cause significant ocular deficits and the necessity of ophthalmological follow-up.

Our study presented various management strategies for orbital incidentalomas, including observation, surgery, or diagnostic biopsy, depending on factors such as clinical symptoms, tumor size, location, radiologic diagnosis, and surgical risks (15, 16). In our study, one-third of patients underwent surgery or biopsy for orbital incidentalomas. The decision to proceed with surgery was primarily based on concerns about potential malignancy, the presence and severity of ophthalmological signs, and an assessment of surgical risks. We performed an incisional biopsy to differentiate malignancy in one case with an irregular tumor shape observed on imaging, and pathological results confirmed lymphoma. For radiologically benign lesions, the surgery group was significantly more likely to exhibit ophthalmological signs, including proptosis, peripheral diplopia, hypoglobus, and palpable mass (all p < 0.05). Additionally, 71% of tumors in the surgery group were located anteriorly, while 86% in the observation group were located posteriorly, demonstrating a significant difference (p = 0.001). These findings suggest that the anterior orbital incidentalomas are more likely to cause functional or cosmetic deficits but pose better surgical feasibility and lower complication risks. Tumor size was larger in the surgery group compared to the observation group but without statistical significance (p = 0.072), and no significant difference in tumor position (intraconal vs extraconal) was observed. These results suggest that the severity of ophthalmological signs, patient perceptions, and surgical feasibility are more important factors than tumor size in the surgery decision-making process.

In our study, two-thirds of patients were managed without surgery for a median follow-up of 2.77 years and showed no development of new functional deficits. All patients in the observation group declined surgical excision of the incidentaloma as they were not bothered by the ophthalmological signs such as proptosis and peripheral diplopia. Most cases (25/29, 86%) had posterior orbital lesions, bearing more potential surgical risks. During the median follow-up period, most lesions (28/29, 96.6%) remained stable in size and morphology, with two cases showing a decrease in size. Only one CVM case showed minimal growth during follow-up without any functional deficits. These findings highlight that conservative management is often appropriate for orbital incidentalomas rather than surgical removal, particularly for posterior lesions without significant signs.

Our study had several limitations. First, the retrospective design, small sample size, and variation in follow-up duration limit our findings’ generalisability. Nevertheless, this study represents the first retrospective review of orbital incidentalomas, highlighting the need for further research to validate and expand upon these initial observations. Second, two cases where tumor removal was expected to yield favourable outcomes were not treated, as the patients declined surgery. This may have affected our ability to evaluate comparisons between the surgery and observation groups. The lack of a standardised protocol for managing orbital incidentalomas resulted in treatment decisions being influenced by individual clinicians’ judgments and patients’ preferences, potentially introducing variability in management approaches. In this study, we adopted a practical approach based on our clinical experience and comprehensive counselling with each patient regarding potential surgical complications before finalising their decisions. Importantly, no adverse clinical outcomes were observed in patients who underwent surgery or those who opted for conservative management.

In conclusion, our findings suggest that orbital incidentalomas are often benign and asymptomatic, and their prevalence is expected to increase due to the growing demand for health check-ups and neuroimaging. Most orbital incidentalomas remain stable in size over time and typically do not require surgery. Observation proves to be a practical management strategy for asymptomatic lesions, particularly those located posteriorly and intraconally. Surgical intervention should be reserved for symptomatic lesions or those located anteriorly, where safe excision is more feasible. Future studies are needed to assess the incidence and establish specific guidelines for the management and follow-up of orbital incidentalomas based on their individual characteristics.

## Data Availability

Our data contain potentially identifying information. Data are available from the Asan Medical Center Institutional Data Access / Ethics Committee (contact via) for researchers who meet the criteria for access to confidential data.

## Author contributions

**Conceptualization:** Ho-Seok Sa.

**Data curation:** Yeong A Choi, Min Kyu Yang, Ho-Seok Sa.

**Formal analysis:** Yeong A Choi, Min Kyu Yang.

**Supervision:** Ho-Seok Sa.

**Validation:** Ho-Seok Sa.

**Writing – original draft:** Yeong A Choi, Min Kyu Yang.

**Writing – review & editing:** Ho-Seok Sa.

## Notes

### Competing Interest Statement

The authors have declared no competing interest.

### Funding Statement

The author(s) received no specific funding for this work.

### Author Declarations

This research was approved by the Institutional Review Board (IRB) of Asan Medical Center (Seoul, Korea IRB number: 2024-1462) and complied with the Health Insurance Portability and Accountability Act.

